# Feasibility study to assess the delivery of a novel isometric exercise intervention for people with high blood pressure in a healthcare setting

**DOI:** 10.1101/2024.02.16.24302961

**Authors:** Jonathan Wiles, Ellie Santer, Melanie Rees-Roberts, Rachel Borthwick, Timothy Doulton, Pauline A. Swift, Tracy Pellatt-Higgins, Katie Saxby, Ashley Mills, Katerina Gousia, Douglas MacInnes, Jamie M. O’Driscoll, Alan West, John Darby, Vanessa Short, Christopher K. Farmer

## Abstract

**BACKGROUND:** Hypertension affects 1 in 4 adults and increases the risk of CV disease. Management aims to reduce blood pressure to a level that minimises risk; however, up to 50% of people fail to achieve blood pressure targets often due to insufficient treatment or poor adherence. Exercise has a role to play in the management of hypertension. The impact of isometric exercise on hypertension in healthcare settings is poorly understood.

**METHODS:** Randomized controlled open label multicentre feasibility study of isometric exercise compared to standard care in unmedicated hypertensives. Participants received an individualized isometric wall squat prescription and performed 4 x 2-minute bouts thrice weekly for 6-months. We assessed recruitment, deliverability, attrition, adherence, and variance in blood pressure change.

**RESULTS:** 41 participants (56 ^+^/- 15 years), 59% women, were randomized. Isometric exercise was found to be easily deliverable to all participants. At 6-months 34% withdrew, of those who completed isometric exercise 87% of their sessions were at the correct intensity. Variance in blood pressure change was 14.4 mmHg. The study was not powered to show a difference in blood pressure between groups, however blood pressure reductions were seen in the intervention group at all study time points compared to baseline.

**CONCLUSIONS:** The results have allowed us to calculate a sample size (n=542) for a full randomised controlled trial. The results demonstrate good acceptability and adherence rates to the treatment protocol. Our results show a signal towards a consistent systolic blood pressure reduction in the isometric exercise group compared to baseline.

**REGISTRATION:** Trial number: NCT04936022

https://classic.clinicaltrials.gov/ct2/show/NCT04936022?cond=isometric+exercise&draw=2&rank=7

**Registry Identifier:** ISRCTN 13472393

## Introduction

Hypertension is very common with a global age-standardized prevalence in adults aged 30–79 years of around 32% in women and 34% in men^1^. The Framingham Heart Study demonstrated that systolic blood pressure (sBP) increases linearly with age after 30 years in industrialized populations^2^. In 2019, the leading Level 2 risk factor globally for attributable deaths was high sBP, accounting for 10·8 million deaths^3^. A major concern to patients and health providers is that only 30% of patients with high BP are being treated effectively^4^. Of those on anti-hypertensive therapy up to 50% of people fail to achieve their target mainly due to non-adherence (estimated 30%-50% failing to adhere)^5, 6^, with undesirable side effects of antihypertensive medication often cited in this context^7, 8^. Mennini et al.^9^ estimated the direct cost associated with hypertension in Europe is €51.3 billion ($55.8 billion). Moreover, the human cost is immense with people suffering significant morbidity from resultant cardiovascular disease. The global population is ageing and although other factors such as improved dietary habits may attenuate risk, the prevalence of hypertension will still increase.

The importance of lifestyle changes for patients with hypertension in the absence of other risk factors should not be overlooked^10, 11^. Exercise has anti-hypertensive benefits^12, 13, 14^ and may be as effective as medication in controlling BP^15^. However, low adoption and high attrition rates are common^16, 17^. Indeed, worldwide, around a third of women and a quarter of men do not do enough physical activity to stay healthy^18^. It is noteworthy that guidance in relation to exercise is generic with the same recommendations for those with and without hypertension^18^. There are also data to suggest that people with hypertension are less physically active than those without hypertension^19^. To exacerbate this situation further, a recent study showed only one in five general practitioners (GPs) are broadly or very familiar with national physical activity guidelines and as many as 72% of GPs do not speak about the benefits of physical activity to their patients^20^. To promote lifestyle exercise changes, people need easy, effective and manageable exercise interventions as a first line option for managing their BP.

Current physical activity guidelines prioritise at least 150-minutes of moderate intensity or 75-minutes of vigorous intensity aerobic activity a week. However, research now acknowledges the importance of frequently cited barriers to exercise such as lack of time and resources^21^, which may help to explain the poor adherence (67%) and high attrition rates (50%) to this relatively large amount of aerobic exercise often reported^16^.

Isometric exercise (IE) training has been consistently shown to reduce clinic and ambulatory BP in both sexes^22^. Only 24-minutes of IE each week are required to achieve reductions in BP of 12/6 mmHg in unmedicated hypertensives, which can be easily performed at home without costly equipment^23^. Moreover, large scale pairwise and network meta-analysis clearly demonstrates that this type of exercise is the most effective exercise mode to reduce sBP when compared to other forms (including moderate intensity aerobic) and combinations of exercise training^24^. Whilst IE may provide a viable solution for those with uncomplicated hypertension, evidence for the effectiveness of IE in this clinical population is still not robust. At present, an IE training intervention has never been tested within a national healthcare setting, nor confirmed in any UK community-based randomized control trials. As such, this feasibility study aimed to inform the design of a large-scale randomized controlled trial to evaluate the efficacy and mechanisms of wall squat IE to lower BP in UK National Health Service (NHS) patients who have sBP between 140-159mmHg not taking antihypertensive treatment^25^.

## Aim

To determine the feasibility of delivering a personalized isometric exercise intervention for people with high BP.

### Primary Outcomes

- To determine the variance in BP change, to enable a sample size calculation for a randomized controlled trial.
- To assess if nurses/allied healthcare professionals can deliver IE prescriptions.

### Secondary Outcomes

- Evidence the fidelity of the study intervention with respect to patient completion of IE.
- Determine short- (4-weeks), medium- (3-months) and long-term (6-months) adherence rates to IE.
- Determine recruitment and attrition rates.

## Methods

### Study design

This study was a multi-centre randomized controlled feasibility trial (RCT) of an IE intervention for patients with sBP 140-159mmHg not taking antihypertensive treatment, carried out in four primary and secondary (added due to logistical difficulties with delivery due to the Covid-19 pandemic) health care sites in the south-east of England.

Patients were excluded if they were taking anti-hypertensive medication; average home systolic BP <135 mmHg; were unable to undertake the study intervention; had a previous history of diabetes mellitus, ischaemic heart disease, moderate or severe stenotic or regurgitant heart valve disease, atrial or ventricular arrhythmia, stroke or transient ischaemic attack, aortic aneurysm, peripheral arterial disease, uncorrected congenital or inherited heart condition; had an estimated glomerular filtration rate <45 ml/min; had a documented left ventricular ejection fraction <45% or left ventricular hypertrophy; had a documented urine albumin to creatinine ratio >3.5 mg/mmol; were unable to provide informed consent; were enrolled in another clinical trial; were pregnant or currently breast feeding; had any medical condition that would make the participant unsuitable for the study.

All procedures conformed to the Declaration of Helsinki principles, and the National Research Ethics Committee approved the study (REC ref: 20/LO/0422, IRAS ID: 274676). Signed informed written consent was obtained from all participants.

Participants agreed to four remote study appointments, one follow-up telephone call, and an additional in-person visit for those allocated to the IE training groups. The appointments comprised a screening visit (Day -7), baseline assessment (Visit 1, Day 1), follow-up visits at 4-weeks (Visit 2), 3- months (Visit 3) and 6-months (Visit 4). A follow-up telephone call was conducted at 1-week (Day 7- 10) to check how the participant was coping with their new exercise programme and to collect heart rate (HR) and BP data. In addition, participants completed questionnaires on diet, exercise and quality of life at each visit. At the end of the follow up period, all participants returned to standard care and control participants were offered an IE training programme (see Figure 1).

**Figure 1.**
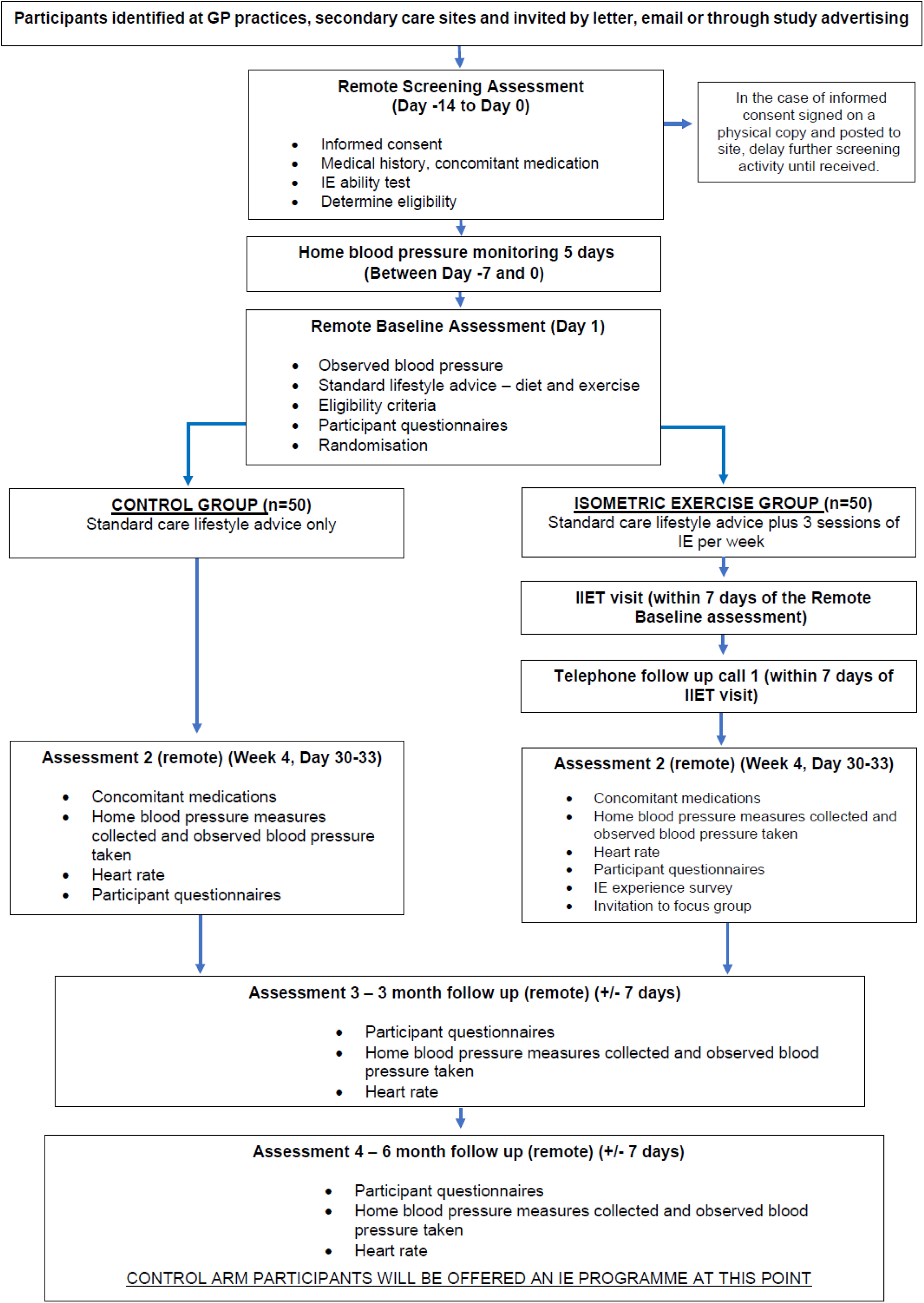
Trial flow chart GP – General Practitioner, IE – Isometric Exercise, IIET – Incremental Isometric Exercise Test

**Figure 2:**
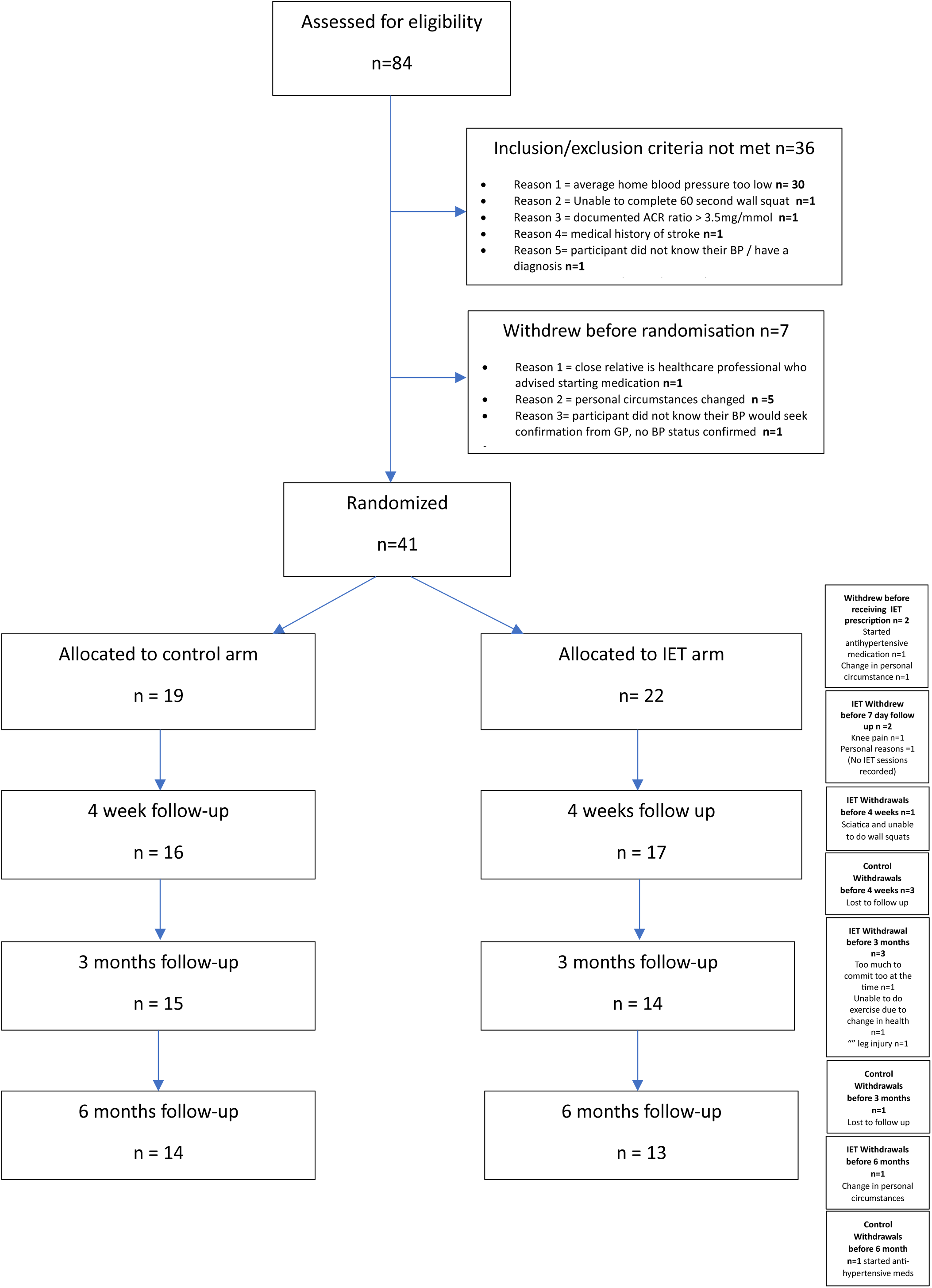
Consolidated Standards of Reporting Trials (Consort) flow diagram illustrating recruitment and follow-up

### Participants

The study design aimed to recruit through primary care; however, our study opened at the beginning of the COVID-19 pandemic and as a result alterations had to be made to the study protocol and design. As discussed by Farmer et al. (2023)^26^, we shifted our focus to direct-to-public advertising through posters at community venues (pharmacies, supermarkets, places of work) and to social media advertising whilst still supporting NHS sites to recruit where possible. Overall, 84 participants consented to enter the study, 7 withdrew prior to screening and 36 were screen failures. In total 17 male and 24 female patients (aged 56.6 ± 14.6 yrs) with a screening sBP between 140-159 mmHg, not yet on anti-hypertensive medication and without any significant medical conditions (See table 1) were recruited.

**Table 1:** Demographic profile of participants.

|  | **Control** | **Isometric Exercise** |
| --- | --- | --- |
|  | **n=19** | **n=22** |
| Male n (%) | 8 (42.1) | 9 (40.9) |
| Age (years), mean (SD) | 56.2 (14.3) | 57.0 (15.2) |
| Age ≥ 50 years, n (%) | 14 (73.7) | 16 (72.7) |
| Height (cm), mean (SD) | 170.3 (12.1) | 171.0 (9.9) |
| Weight (kg), mean (SD) | 80.7 (15.5) | 79.1 (15.1) |
| BMI (kg/m<sup>2</sup>) | 27.7 (4.5) | 26.9 (4.2) |
| Smoking status n (%) |  |  |
| Smoker | 1 (5.3) | 2 (9.1) |
| Previous smoker | 8 (42.1) | 6 (27.3) |
| Non-smoker | 10 (52.6) | 14 (63.6) |

### Procedures

Following the initial remote screening visit, eligible patients were sent a home BP monitor (Omron M3 Intellisense, Omron Electronics Ltd., UK). They received instructions (verbal, written and video) on how to use this following British and Irish Hypertension guidelines (http://bihsoc.org/wp-content/uploads/2017/11/BP-Measurement-Poster-Automated-2017.pdf) and took three readings to confirm their BP status. Initially participants were asked to report their body weight and height. At the end of this period eligible patients were then randomized in a 1:1 ratio to receive either an IE programme in addition to standard care advice or continue with standard care advice alone for 6- months. We subsequently collected weight by questionnaire at each time point in both intervention and control groups (see Figure 1). At each study (virtual) visit, BP measurement was recorded using video-supervised home BP readings (as opposed to unsupervised).

### Intervention description

Standard care lifestyle advice (Group 1 – control) or wall squat IE training with standard care lifestyle advice (Group 2).

The standard care lifestyle advice provided to all participants taking part in the study included: recommended daily salt intake of 5 to 6 g/day; healthy diet rich in fresh fruit and vegetables and low in saturated fat; maintain a body mass index (BMI) between 20 and 25 kg/m2; keep alcohol consumption to less than 14 units (10 US units) per week; exercise to make you breathless for at least 30 min five times a week.

For those allocated to the IE group an additional in-person visit took place within 7-days of the remote baseline assessment. The participants carried out an incremental isometric exercise test (IIET), the results of which were used to prescribe the correct individual wall squat intensity for future IE training sessions conducted in the home. This was determined by the knee joint angle required to elicit a target HR of 95% HR_peak_. Target heart rate range (THRR) was established for each participant to ensure future IE training sessions were of the required intensity^27^. At the end of this session, the participant was taken through the necessary information to successfully complete the wall squat IE in their home and to ensure that they fully understood what was required of them throughout the 6-month training period (for full protocol see Wiles et al.^25^). All IE training sessions thereafter were completed in the home. Each IE training session comprised of four bouts of 2-minute wall squats with 2-minutes recovery in-between, with HR recorded at the end of each bout. Participants were asked to perform three IE training sessions a week, ideally on alternate days to allow for adequate between session recovery.

Participants received reminders (texts/email) to encourage adherence and collect home BP measurements. The control group received monthly reminders to adhere to the standard care lifestyle advice given by their Health Care Professional (HCP) at baseline. The intervention group also received monthly standard care lifestyle advice in addition to three IE training reminders per week, for the duration of the study. At follow up timepoints 4-weeks, 3-months and 6-months, all participants received a 24-hour reminder to start taking their home BP.

### Data Analysis

Data were analysed based on intention-to-treat (ITT) for all patients randomized, and per protocol (PP). Participants in the intervention group who completed at least eight of 12 exercise sessions between baseline and the 4-week timepoint were included in the PP dataset, all participants in the control group were also included.

The primary and secondary outcomes for sBP, diastolic BP (dBP) and HR were analysed using a mixed model with repeated measures over time, and an unstructured covariance matrix. The model included a fixed treatment effect to compare change from baseline between IE and control groups, and adjustment was made for baseline values, sex and age (18-49, ≥50 years). This model was used to estimate differences between the treatment groups and 95% confidence intervals. Assessment of residuals and diagnostic plots supported the assumptions of normality and it was not necessary to perform data transformations prior to analysis.

Outcomes, demographic and baseline data were summarized to compare treatment groups. Means and standard deviations were calculated for continuous (approximate) normally distributed variables, and frequencies and percentages for categorical variables.

The quantitative analysis was conducted using RStudio version 2023.06.1 and Stata/IC 16.1. For the economic analysis, we used the EQ-5D-5L questionnaire to measure health status at each assessment point. The responses were converted into utility scores using the EQ-5D-5L value set for England^28^. Resource use was collected using the Client Service Resource Inventory (CSRI). We adopted the UK NHS plus PSS (personal social services) perspective. Data were undiscounted due the shorter follow-up (up to 6-months post-randomisation). Unit costs of health and social care^29^ were applied to obtain individual service use data. Costs were calculated per participant in each group. Stata 17 and Microsoft Excel (2019) were used to perform the analysis.The cost elements for delivering the IE intervention comprised of training of clinical staff (research staff costs), clinical staff costs to deliver the intervention in a healthcare setting and equipment. Equipment to deliver the intervention consisted of a carboard wall squat delivery device, the HR monitor and the BP monitor.

## Results

This RCT met all the progression criteria except the recruitment target, mainly due to the COVID pandemic or its effects. However, there were multiple adaptations implemented to mitigate this^26^. More importantly, this investigation was able to show that IE is acceptable to NHS patients/healthcare professionals.

Statistical summaries of the demographic data indicated that characteristics were very similar between groups. Similarly, there was no indication of differences between groups in the continuous outcomes, sBP, dBP and HR at baseline, weight and BMI (table 1 and 2).

Of the 41 participants randomized, 14 participants (34%) withdrew or were lost to follow-up during the study, 9 participants in the intervention group and 5 participants in the control group. Of note, two participants in the IE group withdrew between randomisation and IE prescription for reasons unrelated to the intervention (summary statistics are outlined in Table 2).

**Table 2:** Summary statistics of blood pressure and heart rate at baseline and over time by treatment group.

|  | Time | Control | Isometric Exercise |
| --- | --- | --- | --- |
| Systolic BP (mmHg) | Baseline | 153.7 (13.2) | 152.3 (12.3) |
| Mean (SD) | 4-weeks | 150.4 (13.8) | 147.2 (14.6) |
|  | 3-months | 150.6 (8.3) | 145.2 (10.1) |
|  | 6-months | 145.4 (12.9) | 142.4 (8.1) |
| Diastolic BP (mmHg) | Baseline | 89.7 (7.3) | 94.0 (8.8) |
| Mean (SD) | 4-weeks | 92.3 (8.4) | 92.4 (10.0) |
|  | 3-months | 92.0 (7.5) | 95.8 (17.3) |
|  | 6-months | 88.5 (8.1) | 89.9 (7.8) |
| Heart rate (bpm) | Baseline | 76.7 (13.4) | 71.3 (8.8) |
| Mean (SD) | 4-weeks | 68.5 (11.0) | 73.0 (11.1) |
|  | 3-months | 69.4 (11.2) | 71.5 (8.07) |
|  | 6-months | 66.8 (11.4) | 69.0 (9.2) |

In the PP analysis for the primary outcome, change from baseline in sBP, the difference in adjusted means between groups at 4-weeks was -3.61 mmHg. sBP reduced in both groups at 4-weeks compared to baseline, but a larger reduction was seen in the intervention group (group means -8.48 mmHg and -4.86 mmHg for the intervention and control group respectively). Similar results were seen at 3-months and 6-months, the differences in adjusted means between groups were -4.01 mmHg and -2.7 mmHg respectively (see table 3.1). ITT analysis supported the PP results, the between group difference in adjusted means of sBP change from baseline was -2.3 mmHg at 4-weeks, -4.07 mmHg at 3-months, and -2.3 mmHg at 6-months (see table 3.2).

**Table 3.1:** Estimates of treatment differences from ANCOVA (Per Protocol population)

|  | Change from<br>baseline | Isometric Ex<br>adj. mean | Control<br>adj. mean | Difference adj.<br>mean | Difference<br>95% CI |
| --- | --- | --- | --- | --- | --- |
| Systolic BP | 4-weeks | -8.48 | -4.86 | -3.61 | -13.08 to 5.86 |
| (mmHg) | 3-months | -8.17 | -4.15 | -4.01 | -10.57 to 2.54 |
|  | 6-months | -12.87 | -10.20 | -2.67 | -12.65 to 7.31 |
| Diastolic BP | 4-weeks | -1.07 | 0.89 | -1.96 | -7.60 to 3.68 |
| (mmHg) | 3-months | 2.78 | 0.88 | 1.90 | -8.90 to 12.70 |
|  | 6-months | -1.45 | -2.28 | 0.82 | -5.36 to 7.00 |
| Heart rate | 4-weeks | 0.09 | -6.10 | 6.19 | -1.45 to 13.83 |
| (bpm) | 3-months | -1.90 | -6.38 | 4.49 | -3.84 to 12.81 |
|  | 6-months | -2.92 | -8.80 | 5.88 | -3.15 to 14.92 |

Adj. – Adjusted; CI – Confidence Interval

**Table 3.2:** Estimates of treatment differences from ANCOVA (Intention-to-treat population)

|  | Change from<br>baseline | Isometric Ex<br>adj. mean | Control<br>adj. mean | Difference adj.<br>mean | Difference<br>95% CI |
| --- | --- | --- | --- | --- | --- |
| Systolic BP | 4-weeks | -6.42 | -4.11 | -2.31 | -11.29 to 6.67 |
| (mmHg) | 3-months | -7.58 | -3.52 | -4.07 | -10.28 to 2.15 |
|  | 6-months | -11.92 | -9.56 | -2.35 | -11.70 to 6.99 |
| Diastolic BP | 4-weeks | -0.31 | 1.24 | -1.55 | -6.77 to 3.66 |
| (mmHg) | 3-months | 2.66 | 1.15 | 1.51 | -7.73 to 10.75 |
|  | 6-months | -1.23 | -1.99 | 0.76 | -5.40 to 6.91 |
| Heart rate | 4-weeks | 0.04 | -5.57 | 5.61 | -1.10 to 12.31 |
| (bpm) | 3-months | -1.34 | -5.96 | 4.62 | -2.86 to 12.10 |
|  | 6-months | -1.44 | -8.34 | 6.90 | -0.81 to 14.61 |

Adj. – Adjusted; CI – Confidence Interval

Analysis of dBP and HR did not show a similar pattern; there was little evidence of any change for either of these outcomes (Tables 3.1 and 3.2).

As expected, the study did not detect a difference between the control and intervention group at any time point, because it was not powered to. However, post-hoc PP (and ITT) analysis investigating the difference in sBP between baseline and each timepoint suggested a reduction in sBP (based on CI) at all three timepoints in the IE group, but only a reduction in the control at 6-months (Figure 3, PP analysis).

**Figure 3.**
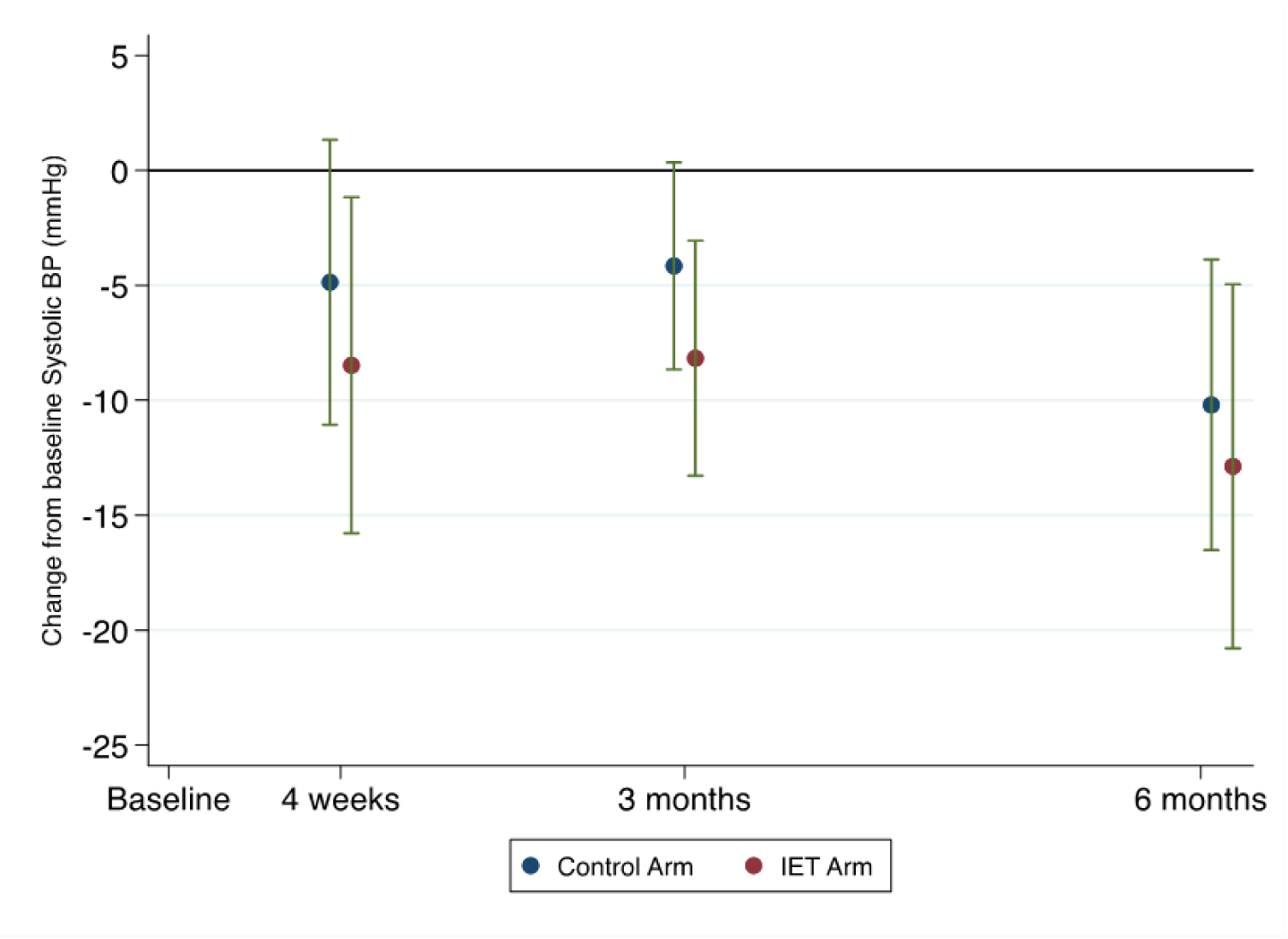
Marginal means and 95% confidence interval from ANCOVA (Per Protocol population)

There was no trend to reduction in body weight in either group. However, of note were two participants in the control group who had lost a considerable amount of weight at 6-months (8.8kg and 11.9kg).

All HCPs who carried out an IIET with a participant passed the competency assessment.

We measured compliance with IE using the HR responses in bouts 3 and 4 of each session. Compliance was deemed adequate if at least two thirds of HRs were in the participant’s THRR. Thirteen of the IE participants returned HR data throughout the study. At the beginning of the study (1-week) 69% of the 12 training sessions were in range and at 6-months 85% of the 72 sessions were within range.

Estimates of the SD for sBP change from baseline at 4-weeks, 3-months and 6-months are 12.5, 10.8 and 14.4mmHg respectively.

### Health Economic Analysis

Out of the 33 participants who completed an EQ-5D questionnaire, results were analysed from 21 participants who completed it at baseline and at least one more point. There was no difference in quality of life between groups at any time point.

Regarding the feasibility of collecting health economic data, we had completed resource utilization questionnaires for 18 participants at 4-weeks, 19 at 3-months and 14 at 6-months. The changes in resource use between groups and over time were small. Participants did not use any hospital or social care services during the study period but did access primary care services. However, the frequency was very low with no statistical difference between groups. The estimated excess resource use costs for the intervention at 6-months was $0.77 but this was not statistically significant at any time point.

Training and prescription delivery cost was calculated based on hourly wage of different staff involved. The training costs totalled $1,334. The cost of delivering the intervention (performing the IE, calculating the optimal knee angle and adjusting the prescription) totalled $868. Equipment costs outside a research setting were calculated at $949 without the inclusion of the BP monitors and at $2,221 with the inclusion of the BP monitors. The average cost for delivering the intervention outside a research setting was $82.6 per person without including the BP monitor cost and $140 per person including the BP monitor cost.

## Discussion

This is the first study to assess the feasibility of delivering an accessible wall squat IE intervention to NHS patients presenting with hypertension not on anti-hypertensive medication. The results provide the necessary data for sample size calculation for a future large scale RCT of IE training in hypertension. If we accept a minimum clinically important difference in sBP of 5mmHg^24^, using the 6-month standard deviation of 14.4 mmHg, any future studies will require 352 participants to attain 90% power with a 5% statistical significance level. Indeed, this finding concurs with previous expert suggestion that most standard deviation values for change in BPs are above ±14 mmHg and supports the conclusion that previous studies where no differences in BP were found following isometric exercise training may have been underpowered^30^. Thus, based on our finding of a 35% attrition rate, any subsequent RCT would require a total sample size of 542, namely 271 participants in each treatment group.

This study was designed as a feasibility study and was not powered to detect differences in BP. However, it is of interest to note that exploratory post-hoc analysis comparing BP at baseline and subsequent timepoints showed a trend to reduction in BP at all time points in the IE group (ITT = -11.9 mmHg at 6- months). This change in BP is consistent with findings of previous studies looking at effects of 4-weeks and other durations of isometric wall squat training upon BP. Taylor et al.^23^ recruited 24 unmedicated hypertensive males (44 ± 7 years) who were randomly assigned in a crossover study to 4-weeks of isometric wall squat exercise or control. Following the IE training, clinic and 24-h ambulatory BP significantly (P<0.001) reduced by 12 / 6 (± 4) mmHg. The similitude of these mean sBP reductions would also support the findings of Badrov et al.^31^, which demonstrate that IE training lowers BP equally in males and females.

Few studies have investigated the effects of IE training performed over longer durations upon resting BP in hypertensive participants. Two comparable studies examined the effect of isometric handgrip exercise performed at 30% maximal voluntary contraction over 12-weeks^32, 33^ and neither showed significant reductions in office BP following IE compared to control. Whilst there are numerous potential reasons that might explain these findings, arguably a contributing factor may be that not all isometric exercises have the same effect on BP. Indeed, it has been suggested that a potential physiological stimulus for BP adaptation may be linked to post-exercise hypotension^34^. Our research group has previously shown that the wall squat IE has been associated with a greater post exercise hypotensive response than isometric handgrip^35^ and results in a greater magnitude of BP reduction following training^24^. The reality of the situation is most likely reflected in the results of a recent real-world study conducted by Cohen et al.^36^ who compared the effects of 12-weeks of hand grip against wall squat IE and a control group in unmedicated hypertensive participants in a multi-centre trial in Columbia. The study demonstrated significant office sBP reductions of –11.2 mmHg for hand grip and –12.9 mmHg for wall squat groups compared to control (–4 mmHg) but reported a larger magnitude of difference between the wall squat and control. Furthermore, the only study to investigate longitudinal efficacy of IE training as an antihypertensive intervention beyond this duration showed significant reductions (P<0.001) in clinic sBP (-9 mmHg) and dBP (-7 mmHg) compared with control following 1-year of isometric wall squat training in prehypertensive males^37^.

Our results demonstrated that HCPs could deliver the wall squat IE protocol indicating that all participants received an individualized IE prescription. We also demonstrated relatively low attrition at 6-months when compared to (i) adherence to antihypertensive medication, which is estimated to be around 50%^38^ and even lower (around 37%) in younger adults^39^ and (ii) the general finding that around 50% of people who start an exercise program discontinue within 6-months^16^. Our findings suggest that at 6-months, 65% of those prescribed an IE programme continued exercising and were training within the recommended target HR for at least two thirds of their IE sessions. This compares favourably to the findings of Siada et al.^40^ who assessed the long-term adherence of a very similar ‘at-risk’ demographic to a 12-week mixed exercise training programme and found that only 48% of participants met the adherence criteria defined as attending more than 75% of the 24 sessions. Whilst the current study had a higher level of attrition than the 27% reported by O’Driscoll et al.^37^, it must be remembered that the latter was a laboratory-controlled study with direct contact between the researchers and participants as opposed to a real-world study, where following an initial face-to-face IE prescription the exercise programme was delivered entirely remotely. Moreover, the current study recruited a more heterogeneous population rather than university staff and students generally recruited in many of smaller scale lab/home-based studies to date; with the age and gender profile of study participants better reflecting that of the general hypertensive population^41^.

Real world randomized controlled exercise intervention design is extremely difficult to achieve, not least in relation to the control group, who for simplicity of comparison would ideally remain sedentary. However, for the purposes of this investigation mandated physical inactivity would not be recognized as either standard care or an effective clinical option^42^, hence the obligation to reinforce the need for standard care lifestyle advice to the control group (and thus also the intervention group) in this study. Like any RCT in hypertension, a trend for reduction in BP^43^ was also seen in the control group (ITT = - 9.6 mmHg), but notably only at 6-month compared baseline. Both the IE and control groups were given detailed written instructions in relation to lifestyle advice in line with NICE 2019 and AHA^44^ physical activity guidelines for hypertension. It could be suggested that this finding was to be expected based solely upon an inevitable Hawthorne effect, but in this instance exacerbated by the fact that people who volunteer for an exercise study will often undertake exercise if allocated to the control group. This problem of control group contamination is well documented in RCTs aiming at increasing physical activity levels^45^ and may account for a potential control group drift to taking on exercise as per lifestyle guidelines in non-blinded exercise studies such as ours. Moreover, NICE lifestyle guidance for hypertension, although generally poorly adhered to is clearly evidence based. Indeed, the dietary advice and the general exercise recommendation that form the basis of standard care advice, if adhered to, have been consistently proven to reduce BP^46, 24^. Furthermore, it has been shown that using information and communication technology (aspects of which were also utilized in our study) to support hypertension management, can significantly enhance BP control after six months using antihypertensive medication^47^.

However, if the BP trends observed in the current study are indicative of the magnitude of reduction that could be achieved in participants who choose to include exercise as a lifestyle modification, the mean data trends indicate that the greatest BP reduction is likely to occur following isometric wall squat training, which would concur with the recent largescale network metanalysis findings of Edwards et al. (2023)^24^. However, more careful monitoring of compliance of control group activity in any future trial will provide greater insight into this phenomenon and allow for more meaningful interpretation of results when discerning the effects of IE upon resting BP^45^.

The BP findings of this feasibility study lend further support for a large-scale RCT to inform any future changes in physical activity guidelines for the prevention and treatment of hypertension. However, the clinical implementation of IE training as the primary recommended exercise mode in managing BP is becoming increasingly irrefutable^24^. The fact that around 40-50% of people treated for hypertension still fail to achieve their target BP^1^ despite available clinical practise guidelines has led to the recognition that in patients with hypertension, exercise should be individually prescribed based upon initial BP level^48^. Indeed, Hanssen and Pescatello^49^ logically identify that other moderators such as comorbidities along with individual preferences and available resources should all be considered when prescribing exercise to this population. Isometric exercise training if shown to be effective in a large RCT would not be advised as a stand-alone exercise treatment for hypertension, but rather as another tool in the exercise guideline armoury to support individual prescription.

There were no reported adverse events associated with the IE in our study which supports our previous findings^50^; however, the study only included people with little or no comorbidity and uncomplicated hypertension not on treatment. Isometric activity is frequently encountered in activities of daily living along with aerobic exercise. Taken together with lower myocardial work and improved myocardial perfusion during diastole^51^ IE is probably as safe as completing aerobic and/or traditional dynamic resistance training^52, 50^. Others have reported that the incorporation of light IE training has been shown to elicit beneficial effects with very limited adverse events in those with cardiovascular disease^53^.

There were two significant problems with recruitment in primary care. The first was case finding, there were significantly fewer individuals with hypertension who were unmedicated than expected. This was in part because the study recruited during and just after the COVID-19 pandemic where patients were not having BP measured^54^. This was exacerbated by the removal of requirement for healthcare providers to record those diagnosed with high BP, but also many people were commenced on anti- hypertensives early after the diagnosis of stage 1 hypertension at odds with national guidance^26, 55^.

Therefore, future studies should include people on antihypertensive medication (except beta-blockers) and allow direct patient recruitment as this was the most effective way of recruiting in this trial.

The screen fail rate was high in this study (50%), this was because many people, once initially screened, did not subsequently have high enough home BPs to confirm the diagnosis of hypertension despite high office readings. Participants were asked to record home BP readings for 5-days resulting in several screen failures. In addition, the protocol included a long list of exclusion criteria because of possible (albeit unlikely) safety concerns. We have now established that IE is well tolerated and conditions such as diabetes, proteinuria or previous myocardial infarction (more than 3-months prior to enrolment) should not be exclusion criteria for future studies.

We found that direct recruitment through social media was highly effective. This required the provision of sites for the prescription of IE outside primary care. We have subsequently developed a method allowing people to accurately self-prescribe in their own homes^56^.

Overall, it was feasible to collect enough resource use and EQ-5D-5L data to conduct the economic analysis, despite the challenges imposed by the COVID pandemic. The use of self-completed questionnaires for both resource use and health data proved a good way to collect these data. We had a 64% response rate at baseline which dropped at subsequent assessment points. We had the highest number of observations at 3-months and the lowest one at 6-months. The analysis indicated that there was no statistically significant difference in effectiveness and resource utilisation costs between the two trial groups, as the feasibility study did not have the necessary statistical power to find a significant effect. A larger study with a greater sample size is therefore desirable to understand better the size of the effect of the intervention on both outcomes and costs. As this is a feasibility study with a very small sample size, these cost-effectiveness estimates are preliminary and not necessarily indicative of the cost-effectiveness of the intervention.

## Limitations

Several limitations warrant consideration. Firstly, the composition of the study population may impede the generalizability of the findings, as it is evident that the representation of minority ethnic individuals is notably lacking. This underrepresentation raises concerns about the broader applicability of the results, potentially leading to an incomplete understanding of exercise behaviours and outcomes across diverse ethnic backgrounds. Additionally, the lack of comprehensive data collection on social deprivation and other Equality, Diversity, and Inclusion (EDI) parameters further limits generalisability. The absence of such vital variables precludes a thorough examination of how exercise habits and their effects might vary among different socio-economic strata and demographic groups. In addition, volunteers for this type of activity tend to come from a cohort who are interested in their health, and therefore may not be completely representative of the wider population with hypertension.

Furthermore, a methodological limitation arises from the inherent impossibility of blinding exercise studies. The nature of the intervention inherently prevents the concealment of exercise assignments from participants, introducing the potential for bias in reporting and compliance. Consequently, the risk of bias influencing participants’ responses to outcomes cannot be disregarded. In this study the investigators were blinded, however supervising HCPs were not which may have introduced bias. In the planned full RCT all staff collecting results will be blinded.

Finally, the challenge of individuals in the control group independently adopting measures beyond the standard guidance introduces a confounding factor, as participants in the control group may implement alternative interventions or lifestyle modifications independently, thereby blurring the distinction between the intervention and control groups. Such uncontrolled variations in the control group could undermine the ability to isolate the specific effects of the exercise intervention under investigation.

## Conclusions

We report the result of a feasibility study, which has allowed us to calculate the sample size (n=542) for a full RCT of IE for people with hypertension. Our results show a signal towards sBP reduction in the IE group compared to baseline and indicate good acceptability and adherence rates to the treatment protocol. Recruitment and delivery of the exercise regimen was challenging in primary care not least because of the COVID-19 pandemic. Since many of these structural difficulties remain, we have developed a self-prescribed approach for the full trial. Our inclusion and exclusion criteria were very conservative and for a full effectiveness RCT, individuals on single BP agents and those with co- morbidities will be included due to good safety data from this and other studies. The cost of the intervention was minimal at less than $90 per patient, but proposed changes to the delivery protocol will reduce costs further. Should a full effectiveness study demonstrate clinically meaningful reductions in BP it is likely that this would be a cost-effective intervention.

## Data Availability

Anonymised data from the study can be provided on request

